# Uncertainties regarding cerebral palsy diagnosis: opportunities to operationalize the consensus definition

**DOI:** 10.1101/2023.06.29.23292028

**Authors:** Bhooma R Aravamuthan, Darcy L Fehlings, Iona Novak, Paul Gross, Noor Alyasiri, Ann Tilton, Michael Shevell, Michael Fahey, Michael Kruer

## Abstract

**Background and Objectives:** Cerebral palsy (CP), the most common motor disability of childhood, is variably diagnosed. We hypothesized that child neurologists and neurodevelopmentalists, often on the frontlines of CP diagnosis in North America, harbor uncertainties regarding the practical application of the most recent CP consensus definition from 2006.

**Methods:** We conducted a cross-sectional survey of child neurologists and neurodevelopmentalists at the 2022 Child Neurology Society Annual Meeting. Attendees were provided the 2006 CP consensus definition and asked whether they had any uncertainties about the practical application of the definition across four hypothetical clinical vignettes.

**Results:** Of 230 attendees, 164 responded to the closing survey questions (71%). 145/164 (88%) expressed at least one uncertainty regarding the clinical application of the 2006 definition. Overwhelmingly, these areas of uncertainty focused on: 1) Age, both with regards to the minimum age of diagnosis and the maximum age of brain disturbance or motor symptom onset, (67/164, 41%), and 2) Interpretation of the term “non-progressive” (48/164, 29%). The vast majority of respondents (157/164, 96%) answered ‘Yes’ to the question: Do you think we should revise the 2006 consensus definition of CP?

**Discussion:** We propose that the uncertainties we identified could be addressed by operationalizing the 2006 consensus definition to support a more uniform CP diagnosis. To address the most common CP diagnostic uncertainties we identified, we propose 3 points of clarification based on the available literature: 1) Motor symptoms/signs should be present by 2 years old; 2) CP can and should be diagnosed as early as possible, even if activity limitation is not yet present, if motor symptoms/signs can be reasonably predicted to yield activity limitation (e.g. by using standardized examination instruments, Brain MRI, and a suggestive clinical history); and 3) The clinical motor disability phenotype should be non-progressive through 5 years old. We anticipate that operationalizing the 2006 definition of CP in this manner could clarify the uncertainties we identified among child neurologists and neurodevelopmentalists and reduce the diagnostic variability that currently exists.

## Introduction

Cerebral palsy (CP) is the most common cause of childhood motor disability and requires neurologic care across the lifespan.^1, 2^ Multiple publications have informed the CP definition,^3–11^ but the 2006 definition serves as the primary guide for current practice.^11^ This statement defines CP as:

> *A group of permanent disorders of the development of movement and posture causing activity limitation, that are attributed to non-progressive disturbances that occurred in the developing fetal or infant brain. The motor disorders of cerebral palsy are often accompanied by disturbances of sensation, perception, cognition, communication and behaviour, by epilepsy and by secondary musculoskeletal problems*.

Unfortunately, even when guided by the 2006 definition, CP diagnostic practices vary between physicians.^12^ For example, up to 40% of physicians would not diagnose CP in the setting of a genetic etiology, even though multiple studies have now shown that individuals with CP can have a contributing genetic etiology for their symptoms^6, 12–19^ and individuals with CP and their families often prefer to retain a CP diagnosis even after a genetic condition is identified.^20^ Some physicians are also wary of providing CP diagnoses to children less than 2 years old, even though data shows that early CP diagnoses can be accurately provided and are almost universally desired by families.^21–25^

Though the overarching conceptualization of CP outlined in the 2006 definition remains valuable, we hypothesized that diagnostic variability may arise from uncertainties regarding the clinical application of specific aspects of the 2006 definition. We have previously demonstrated that child neurologists and neurodevelopmentalists feel that their training background ideally poises them to facilitate early and accurate CP diagnoses.^26^ Here, we assess how a large cohort of North American child neurologists and neurodevelopmentalists perceive the clarity of the 2006 definition with regards to practical clinical application. We hypothesized they would identify areas of uncertainty that can inform efforts to operationalize the 2006 definition to support standardized clinical practice.

## Methods

### Standard Protocol Approvals, Registrations, and Patient Consents

This project was granted a Human Subjects Research exemption by the Washington University Institutional Review Board (IRB ID# 201910233, 11/04/2019).

### Surveyed population

We surveyed attendees of a 1.5 hour in-person seminar titled “What is CP?” at the 51^st^ Child Neurology Society (CNS) Annual Meeting in Cincinnati, OH, USA on Friday, October 14, 2023, which is the largest meeting of child neurologists and neurodevelopmentalists in the US and Canada. The number of attendees was determined via headcount in the room at the seminar half-way timepoint.

### Survey development and administration

Survey questions were developed via iterative discussions between child neurologists (MCK, MS, AT, BA) and an advocate and parent of a young person with CP (PG).

At the start of the seminar, all attendees were presented with the 2006 definition and then answered a set of online survey questions using Microsoft Forms (Microsoft Corporation,

Redmond, WA). These questions elicited demographic information and any uncertainties they may or may not have regarding the clinical application of the 2006 definition, namely:

- Do you have areas of uncertainty regarding the clinical application of the 2006 consensus definition of CP? (Answer choices: Yes/No)

- (If ‘Yes’) Please describe one main area of uncertainty.

To assess how the 2006 definition might be applied in practice, we asked attendees to consider hypothetical clinical vignettes designed to probe potential areas of diagnostic variability.^12^ Attendees were seated at 48 round tables of up to 10 people per table. The room was divided into quadrants such that each grouping of 12 tables was surveyed about their thoughts on one of four vignettes (Table 1). Each attendee was instructed to first consider their assigned vignette independently for 5 minutes, without discussion with their tablemates, and to answer these three questions accessed via a QR code at their seat:

- Would you diagnose this child with CP? (Answer choices: Yes/No)
- Do you feel that the 2006 CP consensus definition provides clear guidance regarding whether you should give this child a CP diagnosis? (Answer choices: Yes/No)

- (If ‘Yes’) What about the definition do you feel lacks clarity regarding this specific clinical scenario?

**Table 1.** Hypothetical clinical vignettes.

|  |  |
| --- | --- |
| Vignette 1 | You see an 8 year old (yo) boy with spastic diplegia. Birth was at 24 weeks gestation. Brain MRI at 2 yo shows mild periventricular leukomalacia. A repeat “surveillance” brain MRI at 8 yo done at another facility shows diffuse cortical volume loss and more extensive periventricular leukomalacia, which you confirm on your review. He has had no regression and has consistently made developmental gains. |
| Vignette 2 | You see a 2 yo girl with spastic and dystonic quadriplegia following uncomplicated term birth and reportedly normal development until sustaining non-accidental trauma at 14 months. Brain MRI demonstrates diffuse white matter and deep grey matter injury. Since her brain injury at 14 months old, she has had a persistent motor disability without developmental regression. |
| Vignette 3 | You see a 16 yo boy with spastic diplegia. Birth was at term and uncomplicated. Brain MRI is normal. You obtain genetic testing and see that he carries a pathogenic mutation in CTNNB1 which has been associated with spastic diplegia that is progressive in ~40% of those affected. He has had no regression and has consistently made developmental gains. |
| Vignette 4 | You see a 6 mo old girl (corrected) born at 24 weeks gestation. Brain MRI shows R>L periventricular leukomalacia. Exam shows a R hand preference with reaching, L sided hyperreflexia and mild spasticity, and axial hypotonia. She is able to tripod sit and is otherwise meeting milestones. |

For the next 30 minutes, attendees at each table, all of whom had been asked to consider the same vignette, discussed their responses, and attempted to reach an agreement regarding their table’s views of how the 2006 definition applied to their assigned vignette. At the end of this discussion period, each table was asked to summarize 1-3 main discussion points to be shared with the seminar attendees, which were read out to the whole seminar room.

Attendees were then asked the same question they were asked at the start of the seminar regarding whether they had any uncertainties regarding the clinical application of 2006 definition. Finally, they were asked:

- Do you think we should revise the 2006 consensus definition of CP? (Answer choices: Yes/No)

### Qualitative analysis

Open-ended responses were analyzed using a conventional content analysis approach.^27^ We built upon the codebook established previously for assessing CP diagnostic variability across physicians and refined this codebook iteratively to characterize response content.^12^ Two investigators (NA, BA) independently coded all responses. Discrepancies were resolved through consensus discussion.

### Data availability

Anonymized data will be shared by request from any qualified investigator.

## Results

### Attendee numbers and demographics

At the half-way mark of the 1.5 hour seminar, 230 individuals were in attendance. 228/230 individuals (99%) responded to at least 1 of 6 surveys administered during the seminar (opening poll, closing poll, and the four surveys corresponding to each vignette), with 161 responding to all surveys (70% complete response rate). These attendees were predominantly physicians in independent practice (101/161, 65%) in an academic setting (118/161, 73%) whose patient populations were at least 5% comprised of individuals with CP (135/161, 75%) (Table 2).

**Table 2.** Survey respondent demographics and rates of initial uncertainties regarding practical application of the 2006 CP consensus definition.

| **Demographic category** | **N in each category, % of Total respondents (Total=161)** |  | **N <u>with</u> uncertainties in each category, % of each category** |  | **N <u>without</u> uncertainties in each category, % of each category** |  |
| --- | --- | --- | --- | --- | --- | --- |
| **Career stage** |  |  |  |  |  |  |
| Medical student | 10 | 6% | 5 | 50% | 5 | 50% |
| Resident/ Fellow | 34 | 21% | 24 | 71% | 10 | 29% |
| Attending |  |  |  |  |  |  |
| 0 to 5 years | 35 | 22% | 20 | 57% | 14 | 40% |
| 5 to 10 years | 23 | 14% | 12 | 52% | 11 | 48% |
| >10 years | 47 | 29% | 28 | 60% | 19 | 40% |
| Other | 12 | 7% | 7 | 58% | 5 | 42% |
| **% of patients with CP** |  |  |  |  |  |  |
| <5% | 14 | 9% | 11 | 79% | 3 | 21% |
| 5-25% | 77 | 48% | 46 | 61% | 30 | 39% |
| 25-50% | 27 | 17% | 17 | 63% | 10 | 37% |
| 50-75% | 11 | 7% | 6 | 55% | 5 | 45% |
| >75% | 6 | 4% | 4 | 67% | 2 | 33% |
| N/A | 26 | 16% | 12 | 46% | 14 | 54% |
| **Practice setting** |  |  |  |  |  |  |
| Academic | 118 | 73% | 43 | 36% | 75 | 64% |
| Private | 10 | 6% | 4 | 44% | 5 | 56% |
| Both | 14 | 9% | 5 | 36% | 9 | 64% |
| N/A | 19 | 12% | 12 | 63% | 7 | 37% |

### Child neurologists and neurodevelopmentalists had uncertainties regarding the clinical application of the 2006 CP consensus definition

The majority (96/161, 60%) endorsed areas of uncertainty at the start of the seminar regarding the clinical application of the 2006 CP consensus definition (64/161 without uncertainties, 1/161 abstaining) regardless of their stage of training (Table 2). Of the 83 respondents who described these uncertainties, the top three described uncertainties were:

- The meaning of “non-progressive” in the definition (17/83, 20%)
- Accounting for genetic etiologies when diagnosing CP (17/83, 20%)
- The meaning of “developing fetal or infant brain” and the associated age cutoffs (11/83, 13%) (Table 3).

**Table 3.**
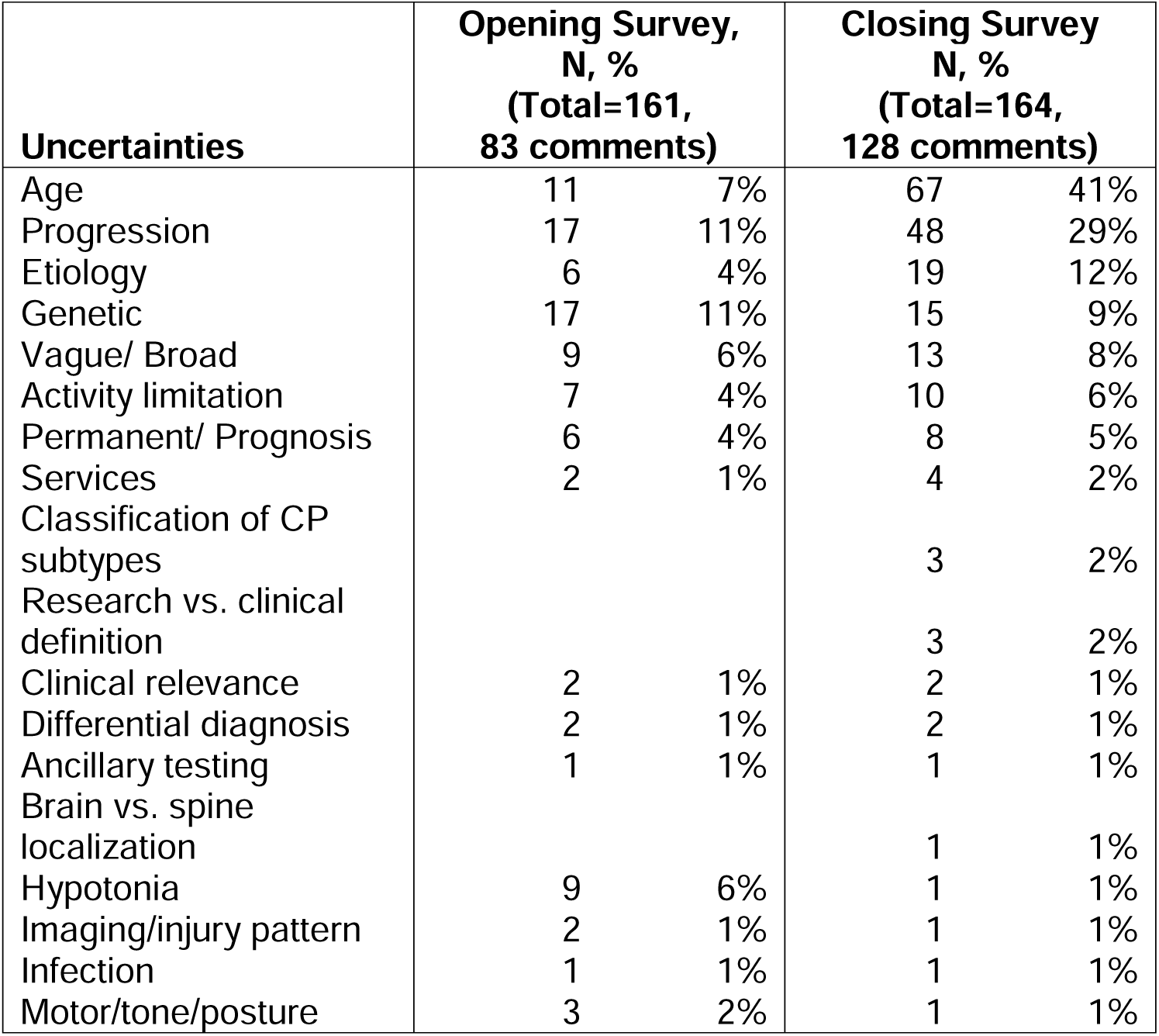
Overall uncertainties regarding the 2006 CP consensus definition at the opening and closing of the seminar.

| **Uncertainties** | **Opening Survey,**<br>**N, %**<br>**(Total=161,**<br>**83 comments)** |  | **Closing Survey**<br>**N, %**<br>**(Total=164,**<br>**128 comments)** |  |
| --- | --- | --- | --- | --- |
| Age | 11 | 7% | 67 | 41% |
| Progression | 17 | 11% | 48 | 29% |
| Etiology | 6 | 4% | 19 | 12% |
| Genetic | 17 | 11% | 15 | 9% |
| Vague/ Broad | 9 | 6% | 13 | 8% |
| Activity limitation | 7 | 4% | 10 | 6% |
| Permanent/ Prognosis | 6 | 4% | 8 | 5% |
| Services | 2 | 1% | 4 | 2% |
| Classification of CP<br>subtypes |  |  | 3 | 2% |
| Research vs. clinical<br>definition |  |  | 3 | 2% |
| Clinical relevance | 2 | 1% | 2 | 1% |
| Differential diagnosis | 2 | 1% | 2 | 1% |
| Ancillary testing | 1 | 1% | 1 | 1% |
| Brain vs. spine<br>localization |  |  | 1 | 1% |
| Hypotonia | 9 | 6% | 1 | 1% |
| Imaging/injury pattern | 2 | 1% | 1 | 1% |
| Infection | 1 | 1% | 1 | 1% |
| Motor/tone/posture | 3 | 2% | 1 | 1% |

Descriptions of uncertainties included:

> *“Resolving possibly progressive genetic diagnoses with ‘non-progressive’ portion of CP definition.”*

> *“The definition of ‘non-progressive’ despite us seeing functional degeneration with time”*

> *“Is there a limit for age of onset and underlying diagnosis?”*

Other described uncertainties included whether to diagnose CP in the setting of pure hypotonia (9/83, 11%), the meaning of “activity limitation” (7/83, 8%), and how etiology should be considered when making a CP diagnosis (6/83, 7%). A minority felt that the entire definition yielded uncertainties (8/83, 10%) with comments including “vague and broad and non-specific” (Table 3).

### The 2006 CP consensus definition did not provide attendees sufficient guidance for how to approach CP diagnosis across four hypothetical vignettes

Across all four vignettes, 75% of attendees (171/228) would diagnose the children in the vignettes with CP while 25% would not. The greatest consensus was for Vignette 1, featuring a child demonstrating radiographic evidence of a potentially progressive brain injury without a progressive phenotype. In this vignette, 93% of attendees would give the child a CP diagnosis. The greatest area of diagnostic variability was for Vignette 3, featuring the child with phenotypically non-progressive spastic diplegia likely due to a pathogenic mutation in CTNNB1, which can result in a progressive phenotype in some. In this vignette, only 54% of attendees would give the child a CP diagnosis (Table 4).

**Table 4.** Perceptions of 2006 CP consensus definition Clarity regarding CP diagnosis across four vignettes.

|  | Would you diagnose CP? |  |  |  | Definition provides clarity? |  |  |  |
| --- | --- | --- | --- | --- | --- | --- | --- | --- |
|  | Yes (N, %) |  | No (N, %) |  | Yes (N, %) |  | No (N, %) |  |
| Vignette 1 (N=43)<br>*(radiographic, not phenotypic, progression)* | 40 | 93% | 3 | 7% | 27 | 63% | 16 | 37% |
| Vignette 2 (N=77)<br>*(traumatic brain injury at 14 months old)* | 54 | 70% | 23 | 30% | 40 | 52% | 37 | 48% |
| Vignette 3 (N=46)<br>*(genetic etiology that can be progressive in some)* | 25 | 54% | 21 | 46% | 15 | 33% | 31 | 67% |
| Vignette 4 (N=62)<br>*(diagnosis of CP in a 6 month old)* | 52 | 84% | 10 | 16% | 41 | 66% | 21 | 34% |
| All (N=228) | 171 | 75% | 57 | 25% | 123 | 54% | 105 | 46% |

Averaged across all four vignettes, 46% of attendees (105/228) felt that the 2006 CP consensus definition did not provide sufficient guidance for how to approach CP diagnosis specific to their assigned vignette. This lack of clarity was again greatest for Vignette 3 featuring the child with CP due a pathogenic mutation in CTNNB1, with 67% of attendees noting that the 2006 definition did not provide clarity regarding CP diagnosis in this situation. Attendees felt the greatest clarity was provided for Vignette 4 featuring a 6 month old child with asymmetric periventricular leukomalacia (PVL) and hemiplegia, but without clear evidence of activity limitation. Yet even for this vignette, 34% of attendees felt that the 2006 definition did not provide clear guidance for how to approach CP diagnosis (Table 4).

Each vignette revealed different areas of uncertainty respondents had about the 2006 definition. For Vignette 1 (CP phenotype with potential radiographically progressive brain injury), the single most cited uncertainty was regarding the interpretation of the term “non-progressive” (cited by 10/43 attendees assessing this vignette, 23%). For Vignette 2 (CP phenotype due to trauma at 14 months old), the most commonly cited uncertainty was regarding the interpretation of “developing fetal and infant brain” (31/77, 40%). For Vignette 3 (CP phenotype due to a genetic etiology that can be progressive in some), the most cited uncertainty was again regarding the interpretation of “non-progressive” (19/46, 41%). Notably, concern about a genetic CP etiology (11/46, 24%) was expressed both because this genetic diagnosis could imply a progressive phenotype and thus obviate a CP diagnosis in the future (6/46, 13%) and because some attendees were unclear whether a genetic etiology inherently precluded a definitive CP diagnosis (5/46, 11%). For Vignette 4 (diagnosis of CP in a 6 month old), a variety of uncertainties about the 2006 definition were noted including the age of diagnosis (7/62, 11%) and the interpretation of “non-progressive” (6/62, 10%), “activity limitation” (6/62, 10%), and “permanent” (5/62, 8%) (Table 5).

**Table 5.**
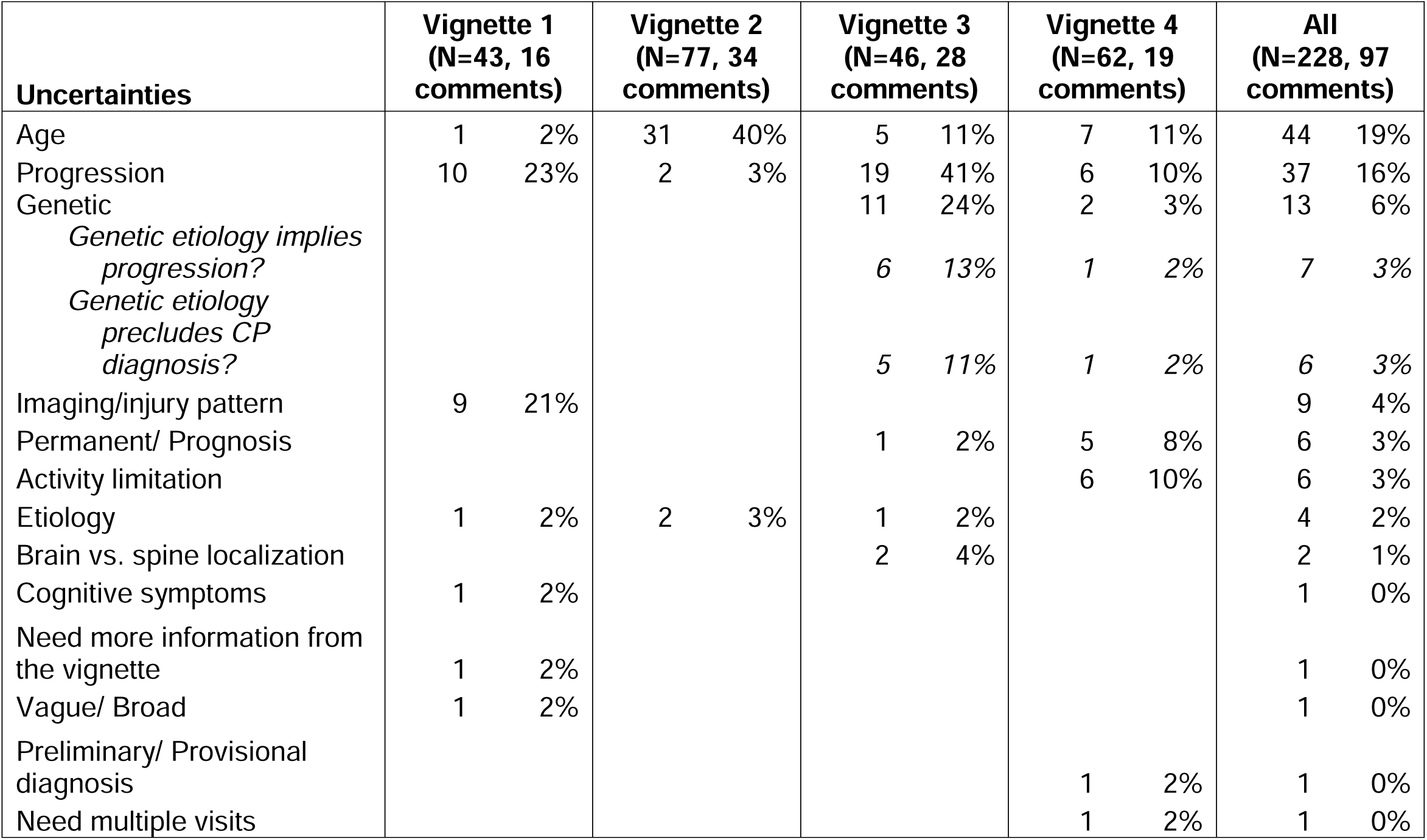
Uncertainties regarding the 2006 CP consensus definition following individual review of each vignette.

| Uncertainties | Vignette 1<br>(N=43, 16<br>comments) |  | Vignette 2<br>(N=77, 34<br>comments) |  | Vignette 3<br>(N=46, 28<br>comments) |  | Vignette 4<br>(N=62, 19<br>comments) |  | All<br>(N=228, 97<br>comments) |  |
| --- | --- | --- | --- | --- | --- | --- | --- | --- | --- | --- |
| Age | 1 | 2% | 31 | 40% | 5 | 11% | 7 | 11% | 44 | 19% |
| Progression | 10 | 23% | 2 | 3% | 19 | 41% | 6 | 10% | 37 | 16% |
| Genetic |  |  |  |  | 11 | 24% | 2 | 3% | 13 | 6% |
| *Genetic etiology implies*<br>*progression?* |  |  |  |  | 6 | 13% | 1 | 2% | 7 | 3% |
| *Genetic etiology*<br>*precludes CP*<br>*diagnosis?* |  |  |  |  | 5 | 11% | 1 | 2% | 6 | 3% |
| Imaging/injury pattern | 9 | 21% |  |  |  |  |  |  | 9 | 4% |
| Permanent/ Prognosis |  |  |  |  | 1 | 2% | 5 | 8% | 6 | 3% |
| Activity limitation |  |  |  |  |  |  | 6 | 10% | 6 | 3% |
| Etiology | 1 | 2% | 2 | 3% | 1 | 2% |  |  | 4 | 2% |
| Brain vs. spine localization |  |  |  |  | 2 | 4% |  |  | 2 | 1% |
| Cognitive symptoms | 1 | 2% |  |  |  |  |  |  | 1 | 0% |
| Need more information from<br>the vignette | 1 | 2% |  |  |  |  |  |  | 1 | 0% |
| Vague/ Broad | 1 | 2% |  |  |  |  |  |  | 1 | 0% |
| Preliminary/ Provisional<br>diagnosis |  |  |  |  |  |  | 1 | 2% | 1 | 0% |
| Need multiple visits |  |  |  |  |  |  | 1 | 2% | 1 | 0% |

Table-based discussion demonstrated comparable areas of uncertainty. Of 41 tables that provided their summary discussion points, 27 (66%) noted uncertainties regarding the interpretation of “developing fetal or infant brain” and the minimum age at which a CP diagnosis can be given and 19 (46%) noted uncertainties regarding interpretation of “non-progressive”. The most commonly mentioned additional point across tables was the importance of a CP diagnosis to qualify for access to necessary services and therapies (22/41 tables, 48%) (Table 6). Example summary statements from these tables included:

**Table 6.** Uncertainties regarding the 2006 CP consensus definition following group discussion by table of each vignette.

| **Uncertainties** | **Vignette 1**<br>**(N=9 tables)** |  | **Vignette 2**<br>**(N=12 tables)** |  | **Vignette 3**<br>**(N=8 tables)** |  | **Vignette 4**<br>**(N=12 tables)** |  | **All**<br>**(N=41 tables)** |  |
| --- | --- | --- | --- | --- | --- | --- | --- | --- | --- | --- |
| Age | 1 | 11% | 12 | 100% | 3 | 38% | 11 | 92% | 27 | 66% |
| Services | 2 | 22% | 9 | 75% | 4 | 50% | 7 | 58% | 22 | 54% |
| Progression | 6 | 67% |  |  | 8 | 100% | 5 | 42% | 19 | 46% |
| Imaging/ Injury pattern | 5 | 56% | 1 | 8% | 1 | 13% | 4 | 33% | 11 | 27% |
| Etiology |  |  | 2 | 17% | 2 | 25% | 4 | 33% | 8 | 20% |
| Genetic | 1 | 11% | 1 | 8% | 4 | 50% | 2 | 17% | 8 | 20% |
| Activity limitation | 1 | 11% |  |  |  |  | 6 | 50% | 7 | 17% |
| Vague/ Broad |  |  | 2 | 17% | 2 | 25% | 2 | 17% | 6 | 15% |
| Permanent/ Prognosis | 1 | 11% | 1 | 8% |  |  | 4 | 33% | 6 | 15% |
| Preliminary/ Provisional diagnosis | 1 | 11% | 1 | 8% |  |  | 2 | 17% | 4 | 10% |
| "CP" term associated with stigma/ Useless | 0 | 0% | 0 | 0% | 1 | 13% | 3 | 25% | 4 | 10% |
| Hypotonia | 2 | 22% | 1 | 8% |  |  | 1 | 8% | 4 | 10% |
| CP due to [etiology] | 0 | 0% | 1 | 8% | 1 | 13% | 1 | 8% | 3 | 7% |
| Brain vs. spine localization | 0 | 0% | 0 | 0% | 3 | 38% |  |  | 3 | 7% |
| Ancillary testing | 1 | 11% |  |  |  |  | 2 | 17% | 3 | 7% |
| Additional diagnosis | 3 | 33% |  |  |  |  |  |  | 3 | 7% |
| Need multiple visits |  |  |  |  |  |  | 2 | 17% | 2 | 5% |
| Misleading families |  |  |  |  |  |  | 2 | 17% | 2 | 5% |
| Motor/tone/posture |  |  |  |  | 1 | 13% | 1 | 8% | 2 | 5% |
| Research vs. clinical definition |  |  | 2 | 17% |  |  |  |  | 2 | 5% |
| Cognitive symptoms | 1 | 11% |  |  |  |  |  |  | 1 | 2% |
| Classification of CP subtypes |  |  | 1 | 8% |  |  |  |  | 1 | 2% |

> *“Re: progression - what about cases where risk of progression is not certain?”*

> *“Need to know age of onset of symptoms - would change our criteria to diagnose CP and not clearly delineated in current definition how to address genetic/potentially progressive or later-onset symptoms.”*

> *“non-progressive - does that mean only clinical or does it include imaging as well?”*

> *“ ‘non progressive’ should refer to a patient’s clinical status, rather than radiologic, though the definition does not specify this”*

> *“avoid repeating imaging without clinical cause”*

> *“Re: definition - when is child no longer ‘infant’ or with a developing brain?” “can you dx at <age 2 years? What about those who improve?”*

> *“Age - confused, how old is too old (fetal/infant - how old)? CP diagnosis gives access but age affects treatment”*

> *“Lower threshold to give a diagnosis of CP if that will provide access to therapies and services”*

*Child neurologists and neurodevelopmentalists feel that the 2006 CP consensus definition should be revised*.

After group discussions, attendees were again asked if they had any uncertainties regarding the 2006 definition. Following 80 minutes of discussion, 145/164 (88%) expressed at least one uncertainty regarding the clinical application of the 2006 definition at the close of the seminar. Overwhelmingly, these areas of uncertainty focused on age (67/164, 41%) and interpretation of the term “non-progressive” (48/164, 29%) (Table 3). Most notably, 157/164 (96%) of respondents responded ‘Yes’ to the question: Do you think we should revise the 2006 consensus definition of CP?

## Discussion

88% of child neurologists and neurodevelopmentalists surveyed expressed uncertainties regarding the clinical application of the 2006 CP consensus definition, particularly concerning age and the interpretation of the term “non-progressive”. Consideration of age yielded uncertainties in two domains: 1) The meaning of “developing fetal or infant brain”, and 2) The minimum age of CP diagnosis, particularly when activity limitation may be contemporaneously absent. 96% felt that the definition should be revised to address these uncertainties.

Attendees did not indicate that any part of the definition was incorrect or redundant, but instead asked for clarification regarding the clinical application of key aspects of the definition. Therefore, explicit clarification of these key aspects may provide an operational version of the 2006 definition that optimizes its clinical utility.

### Why a CP diagnosis matters

Before considering how to operationalize the 2006 definition, it is important to acknowledge the inherent value of a CP diagnosis.

First, a diagnosis of CP has prognostic value. Large registries have demonstrated that children enrolled by age 2 years with a non-progressive motor disability through age 5 years retain a CP phenotype.^5, 28, 29^ Individuals with CP should also be counseled regarding long term risks for associated neurologic and medical conditions.^1, 30^ This prognostication can be valuable medically and also be useful to individuals with CP for long-term planning purposes.

Second, in some parts of the world, a CP diagnosis provides access to therapy services that can yield longstanding benefits if instituted as early as possible.^22–24^ Access to these therapy services is a medical and social justice imperative that the rightful provision of the CP label can unlock.

Finally, a CP diagnosis is firmly desired by those who hold it. Caregivers of people with CP and adults with CP note that a CP diagnosis, more so than any other etiologic descriptor, provides a way to anticipate the evolution of their symptoms, explain their symptoms to others, gain access to services, and become part of a community of people with similar symptoms.^20^ Furthermore, families desire a CP diagnosis as soon as possible and often suspect it long before it is given.^21, 25^

Therefore, operationalizing the 2006 definition should prioritize a prognostically-valuable and inclusive diagnostic paradigm.

### Operationalizing the interpretation of “developing fetal or infant brain”

Disturbances to the developing fetal or infant brain manifest very differently than disturbances to the adult brain. For example, an anatomically identical stroke in an infant versus an adult can differ regarding language involvement, patterns of hemiplegia, risk of epilepsy, and resultant disability.^31, 32^ It is also of note that, though the brain continues to develop and myelinate well into adulthood, most myelination and volumetric growth occurs within the first 2 years of life.^33, 34^

The 2006 definition document explicitly acknowledges uncertainty with regards to the interpretation of “fetal or infant”:

> *“There is no explicit upper age limit specified, although the first two or three years of life are most important in the timing of disturbances resulting in CP. In practical terms, disturbance resulting in CP is presumed to occur before the affected function has developed (e.g. walking, manipulation.).”*

This explanation allows room for interpretation and, therefore, the space for diagnostic variability. For example, 30% of the respondents in this study would not diagnose CP in a child with a permanent motor disability due to traumatic brain injury at 14 months old. Withholding this diagnosis has profound implications for access to medical necessary therapy services, prognostication, and access to a support community. ^20^ To ensure an inclusive and prognostically-valuable operationalization of the 2006 definition, we propose identifying a firm age cut-off by which motor symptom onset (not necessarily disability or activity limitation) must occur to receive a CP diagnosis. Based on the available literature regarding brain development and existing registry enrollment criteria,^5, 29^ we propose an age cut-off of 2 years old by which time motor symptoms must be present to garner a CP diagnosis, supported by psychometrically sound tests.^22–24^

### Operationalizing the minimum age of diagnosis

A CP diagnosis can be reliably provided with high sensitivity and specificity within the first several months of life for an infant with a clinical history putting them at high risk for early life brain disturbance.^22–24^ Early diagnosis is optimized when interpreting standardized neurologic assessments (i.e. the Prechtl General Movements Assessment or Hammersmith Infant Neurological Exam) together with a Brain MRI.^22, 24^ This early diagnosis matters both with regards to management (i.e. earlier institution of therapies is associated with better long term outcomes and prevention of secondary complications) and with regards to providing families some closure in their quest for a diagnostic label. ^21–25^ Yet, practitioners remain reticent to provide early CP diagnoses. Our results show that 16% of attendees would not diagnose CP in a 6 month old child born prematurely with hemiplegia and a supportive pattern of injury on Brain MRI and that 34% feel that the 2006 definition does not provide clear guidance in common this clinical scenario.

We propose that it is important to affirm that a CP diagnosis can reliably be provided at less than 2 years of age in high-risk infants with suggestive standardized exam assessments and Brain MRI even without contemporaneous activity limitation. Instead, if activity limitation is anticipated based on the results of these aggregate assessments, then a CP diagnosis can and should be provided as early as possible.

### Operationalizing the term “non-progressive”

29% of seminar participants were uncertain about how to interpret CP as “non-progressive” in clinical practice.

The concept of radiologic/neurologic progression vs. phenotypic progression was assessed explicitly in Vignette 1. Overwhelmingly, attendees noted that the non-progressive clinical phenotype superseded potentially progressive radiographic brain disease and affirmed that the child in the vignette should be diagnosed with CP. This concept was also explicitly reiterated during table-based discussions: “‘non progressive’ should refer to a patient’s clinical status, rather than radiologic, though the definition does not specify this”.

The 2006 definition clarifies the term “non-progressive” disturbances of the brain as follows:

> *“the pathophysiological mechanisms leading to CP are presumed to arise from a single, <u>inciting event</u> or discrete <u>series of events</u> which are no longer active at the time of diagnosis. This inciting event(s) produce(s) a disruption of normal brain structure and function which may be associated with changing or additional manifestations over time when superimposed on developmental processes. **Motor dysfunction which results from recognized progressive brain disorders is not considered CP.”***

Surprisingly, this explanation contradicts the overwhelming consensus opinion of the seminar attendees. That is, though the 2006 definition applies the word “non-progressive” to “disturbances of the developing fetal or infant brain” and not “activity limitation”, attendees felt the non-progressive activity limitation described in Vignette 1 was enough to give the person a CP diagnosis, despite radiographic evidence of a progressive disturbance. However, diagnosing CP based primarily on a non-progressive phenotype may yield some potential benefits.

First, the mechanisms by which we characterize the progressive nature of the inciting disturbance will evolve with time. For example, some attendees had difficulty reconciling genetic etiologies with a definitively non-progressive condition when the “risk of progression is not certain” (Table 5). Often, little is known about the natural history of rare genetic disorders and the literature is inconsistent in distinguishing between the evolution of symptoms and frank neurodegeneration.^35^ Furthermore, progressive symptoms may only be noted in a subset of affected individuals (e.g. for CTNNB1 as queried in Vignette 3). Although CTNNB1 is now recognized as one of the most common genetic contributors to a CP phenotype, its association with progressive motor disability in some makes it unclear whether it is definitively a “non-progressive” disturbance.^13, 36–39^ Furthermore, as our knowledge of genetic contributors to CP evolves, the genetic contributors that do and do not qualify for a CP diagnosis ideally should not evolve.

Second, it is likely difficult to standardize the definition of “non-progressive” brain disturbances in a resource-responsible and globally-accessible way. In Vignette 1, a brain MRI was financially and medically accessible to the person, despite a lack of true clinical indication (as one table noted: “avoid repeating imaging without clinical cause”). Without the medically unnecessary repeat brain MRI, this child would have unequivocally retained a CP diagnosis. In areas where high-resolution imaging and comprehensive genetic testing are inaccessible, reliance on identifying “non-progressive” brain disturbances will likely result in ongoing diagnostic variability.

Finally, the non-progressive brain disturbance conceptualization of CP should be distinguished from the lifelong functional changes and accumulation of secondary musculoskeletal concerns that occur in individuals with CP. Motor function may decline during young adulthood in individuals with CP, with some losing their ability to ambulate.^40–43^ Many stakeholders are confused by this as they have misinterpreted non-progressive to mean no functional decline.

Therefore, there may be value in limiting the description of “non-progressive” to the childhood period, while simultaneously acknowledging that CP is a permanent condition that evolves across the lifespan.

Supported by large-scale registry data suggesting that re-confirmation of the CP phenotype at 5 years old can be valuable,^5, 28, 29^ we propose that observed or predicted non-progressive motor disability through age 5 should be sufficient for a CP diagnosis. Relative to the 2006 definition, this shifts the application of the term “non-progressive” to “activity limitation” and away from “disturbances”.

The key implication of this shift is that a CP diagnosis should be provided as early as possible without withholding the diagnosis based on uncertainties regarding eventual motor functional decline or improvement in later childhood. That is, if a child has motor symptom onset prior to 2 years old and is either predicted to develop or has developed a non-progressive motor disability (no loss of established motor milestones^44^) through age 5, that child should be given a CP diagnosis. However, if that child goes on to lose motor milestones, this should prompt a diagnostic evaluation for neurodegeneration (likely including brain and/or spine MRI) and possible replacement of their CP diagnosis with a description of the neurodegenerative condition, if applicable. Conversely, if that child does not demonstrate activity limitation due to their motor symptoms in the future, they may have outgrown their CP diagnosis.^28, 45^ This also highlights the importance of screening for CP “mimics” that are treatable (e.g. dopa-responsive dystonia) or neurodegenerative not just at the time of the initial CP diagnosis, but across the lifespan via ongoing motor surveillance.^1, 26^

There may be uneasiness in the medical community that the early provision of a CP diagnosis that is later outgrown or supplanted can cause undue stress for the family.^21, 46^ However, given that families have espoused the value of early CP diagnoses and given the low likelihood of false positive CP diagnoses in the setting of the above criteria, this diagnostic fear may be unfounded.^21, 25, 46^ As has been noted previously, transparency is important when diagnosing CP.^20^ Example language could include:

> *“At this time, your child meets criteria for a CP diagnosis likely due to [etiology(-ies)]. We will continue to track their motor development over time. We do not expect people with CP to lose motor skills in early childhood. If your child ever starts losing motor skills, we will evaluate why and may need to discuss re-evaluating whether or not they still have CP.”*

Or, alternatively, if the child is under 1 year of age:

> *“We expect motor symptoms to cause activity limitation for people with CP. In the future, if your child does not have any motor activity limitations, we will discuss re-evaluating whether or not they still have CP.”*

### Approaches to operationalizing the 2006 CP consensus definition

In summary, we propose four points of clarification to operationalize the 2006 CP consensus definition that are tailored to address diagnostic uncertainties raised by child neurologists and neurodevelopmentalists (Table 7):

1. Motor symptoms or signs should be present by 2 years old^31–34^
2. CP can and should be diagnosed as early as possible, even if activity limitation is not yet present, as long as one can reasonably predict motor symptoms will yield activity limitation (e.g. by using standardized examination instruments, Brain MRI, and a suggestive clinical history)^22–24^
3. The clinical motor disability phenotype should be non-progressive through 5 years old (i.e. no loss of motor milestones ^44^ through 5 years old ^5, 28, 29^)
4. A CP diagnosis should be re-evaluated if there is ever evidence of progressive motor activity limitation or in the absence of motor activity limitation by 5 years of age. ^28, 45^

**Table 7.** Proposed revision to the primary two sentence 2006 CP consensus definition.

|  |  |
| --- | --- |
| **Current wording<sup>11</sup>,**<br>**with proposed**<br>**deletions and**<br>**rearrangements**<br>**<u>underlined</u>** | A group of permanent disorders of the development of movement and posture <u>causing activity limitation</u> that are attributed to <u>non-progressive</u> disturbances that occurred in the developing <u>fetal or infant</u> brain. The motor disorders of cerebral palsy are often accompanied by disturbances of sensation, perception, cognition, communication and behaviour, by epilepsy and by secondary musculoskeletal problems. |
| **Proposed wording,**<br>**with additions**<br>**<u>underlined</u>** | A group of permanent disorders of the development of movement and posture that are attributed to disturbances that occurred in the developing brain <u>causing motor symptom/sign onset prior to 2 years of age<sup>31-34</sup> which can be predicted<sup>22-24</sup> to cause non-progressive activity limitation (no loss of previously established motor milestones<sup>44</sup>) through at least 5 years of age.<sup>5,28,29</sup></u> The motor disorders of cerebral palsy are often accompanied by disturbances of sensation, perception, cognition, communication and behaviour, by epilepsy and by secondary musculoskeletal problems. <u>A cerebral palsy diagnosis should be given as early as possible<sup>21-25</sup> but should be re-evaluated if previously acquired motor milestones<sup>44</sup> are lost or if motor symptoms eventually do not cause activity limitation by 5 years of age.<sup>28,45</sup></u> |

There are many avenues by which these points of clarification can be addressed. Options include revising the primary two sentence description of CP in the 2006 definition (Table 7) or consideration of the 2006 definition as a conceptual one with development of a complementary operational definition for practical use, as has been done for epilepsy.^47^ In addition, new scientific discoveries have been made about CP since the 2006 definition was proposed, including genetic etiological pathways, ^12–18^ clinical practice guidelines for early diagnosis and intervention,^22–24^ and additional pathologies such as an increased risk for cognitive neurodegenerative conditions in adulthood.^48^ A systematic review of this data will likely further inform definition refinement.

### Limitations

We did not assess attendee subspecialties or geographical practice locations, which may provide other sources of variation in CP diagnosis. Approximately ¼ of attendees identified as trainees, though the likelihood of harboring uncertainties regarding the practical application of the 2006 definition did not grossly vary by training status (Table 2). Trainees also add value to this sample population: an operational CP definition should be understandable across training stages.

We surveyed people who chose to attend an in-person seminar on CP at the CNS Annual Meeting. Attendees may have felt they had little CP expertise and were thus attending the seminar to learn or attendees may have had established CP expertise and attended the seminar to discuss the CP definition. Our demographic assessment suggests that people from both ends of this expertise spectrum likely participated in this seminar (Table 2).

Finally, we only present the views of child neurologists and neurodevelopmentalists. However, multiple specialties assume responsibility for diagnosing and caring for people with CP. We propose similar vignette-based sessions within other professional societies to ensure that any uncertainties about the 2006 definition are comprehensively ascertained and addressed. Most importantly, these discussions should engage community stakeholders as a CP definition revision would most directly affect them. We gratefully acknowledge that two authors of this manuscript (PG, NA) are immediate family members of a person with CP.

## Conclusion

Significant practice variation currently exists in the diagnosis of CP. Our findings indicate that much of this variability may be the result of uncertainties regarding the practical clinical application of the current consensus definition of CP. We have elucidated the most common uncertainties held by a large cohort of child neurologists and neurodevelopmentalists. Based on our findings and available evidence, we propose four areas for clarification of the 2006 CP consensus definition. We anticipate that operationalizing the 2006 definition as proposed would reduce clinical practice variation and help standardize the diagnosis of CP.

## Data Availability

Anonymized data will be shared by request from any qualified investigator.

